# Mechanisms and Predictors of Acute Kidney Injury with Perioperative Rosuvastatin in Patients Undergoing Cardiac Surgery

**DOI:** 10.1101/2023.02.09.23285690

**Authors:** RS Wijesurendra, R Sardell, R Jayaram, N Samuel, Z Chen, N Staplin, R Collins, Z Zheng, R Haynes, M Hill, J Emberson, B Casadei

**Author notes:** **Corresponding Author:** Barbara Casadei MD DPhil FRCP FMedSci FESC, **Address:** Division of Cardiovascular Medicine, University of Oxford Level 6, West Wing, John Radcliffe Hospital, Headley Way Oxford, UK, OX3 9DU, **Email:**.

## Abstract

**Background:** In patients undergoing cardiac surgery perioperative statin therapy has been associated with an unexpected increase in postoperative plasma creatinine. Here we investigated mechanisms and predictors of acute kidney injury (AKI) in 1922 patients enrolled in the Statin Therapy in Cardiac Surgery (STICS) randomized placebo-controlled trial of perioperative rosuvastatin (20 mg once daily).

**Methods:** AKI was defined according to international guidelines (KDIGO) using plasma creatinine, and also by cystatin C. Potentially mechanistically relevant plasma/serum biomarkers of muscle injury, inflammation, and kidney injury were investigated, including total creatine kinase (CK), growth differentiation factor 15 (GDF-15), interleukin-6 (IL-6), procalcitonin (PCT), placental growth factor (PLGF), kidney injury molecule-1 (KIM-1), and neutrophil gelatinase-associated lipocalin (NGAL).

**Results:** At 48 hours post-surgery, the incidence of AKI was greater in the rosuvastatin group than in the placebo group when defined by a rise in creatinine (24.7% vs 19.3%, p=0.005) or cystatin C (9.2% vs 5.1%, p<0.001); the majority of AKI was stage 1 in severity (87% when defined by creatinine, and 80% when defined by cystatin C). Compared with placebo, rosuvastatin led to higher postoperative serum levels of KIM-1 (278±5 pg/ml versus 259±5 pg/ml, P=0.01), and to more frequent elevations in CK to >10x and >40x the baseline level (30.9% versus 26.5%, p=0.032, and 2.1% versus 0.7%, p=0.016, respectively), whereas postoperative concentrations of GDF-15, IL-6, PCT, PLGF, and NGAL were similar between groups. In multivariable analyses, insulin treatment, baseline KIM-1, combined coronary artery bypass grafting (CABG) and aortic valve replacement (AVR) surgery, and allocation to rosuvastatin were all independently associated with AKI as defined by creatinine or cystatin C. Odds ratios for rosuvastatin compared to placebo for both creatinine- and cystatin C-defined AKI were not materially altered by further adjustment for post-randomization increases in CK.

**Conclusions:** Perioperative rosuvastatin initiation increased the absolute risk of AKI after cardiac surgery by 4-5%, whether defined by creatinine or cystatin C, and led to higher post-operative KIM-1, suggesting a deleterious effect on renal function, possibly mediated by proximal tubular injury. Insulin treatment, baseline KIM-1, combined CABG/AVR surgery, and allocation to rosuvastatin were all independently associated with AKI by any definition.

## Introduction

Statins are one of the most widely used classes of prescription medicine across the world^1, 2^. The efficacy and safety of statins have been robustly demonstrated^3^, with relative reductions in rates of major cardiovascular events proportional to the absolute LDL cholesterol reduction^4^. In addition to reducing synthesis of cholesterol by competitive inhibition of HMG-CoA reductase, various pleiotropic effects of statins, including anti-inflammatory^5^ and antioxidant^6, 7^ effects, have also been described. As a result, statins have been proposed as a potential modulator of clinical outcomes after surgery^8^. In particular, some observational studies suggested that statins may be associated with a reduced risk of acute kidney injury (AKI) after cardiac surgery^9^, although this has not been a consistent finding^10, 11^. Any effect on postoperative AKI is relevant as this complication affects as many as 25-30% of patients after cardiac surgery^12, 13^ and is associated with longer hospital stay and increased risk of mortality^14-16^.

The Statin Therapy In Cardiac Surgery (STICS) trial was a randomized double-blind placebo-controlled trial that aimed to provide definitive evidence regarding the effects of perioperative statin therapy on the co-primary outcomes of postoperative atrial fibrillation and perioperative cardiac injury in patients undergoing elective cardiac surgery^17^. Whilst perioperative rosuvastatin 20 mg once daily did not significantly affect either co-primary outcome, AKI defined by Acute Kidney Injury Network criteria^18^ (a pre-specified secondary outcome) was unexpectedly found to be significantly more common in rosuvastatin allocated patients^17^.

In addition to STICS, a smaller trial of high-dose perioperative atorvastatin in patients undergoing cardiac surgery did not support the initiation of statin therapy to prevent postoperative AKI after showing a null result and being stopped prematurely after 615 patients had been randomized^19^. Other studies investigating the incidence of AKI in patients treated with statins compared to control therapy after cardiac surgery have reported variable results, but these were all small (≤200 patients each) and many had additional limitations^20-28^.

To understand the mechanism and predictors of post-operative AKI in patients undergoing cardiac surgery treated with perioperative rosuvastatin, we undertook further analysis of the STICS data and samples, including measurement of cystatin C as well as analysis of several biomarkers relevant to muscle injury, inflammation, and kidney injury.

## Methods

### Trial design

The methodology and primary results of the STICS trial (NCT01573143) have been published previously^17^. Briefly, STICS was a randomized double-blind placebo-controlled trial of perioperative rosuvastatin in 1922 Chinese patients undergoing elective cardiac surgery. Men and women who were 18 years of age or older and were scheduled to undergo elective coronary-artery bypass grafting (CABG), surgical aortic-valve replacement (AVR), or both were eligible if they were in sinus rhythm and were not taking antiarrhythmic medication (other than beta-blockers). Patients were excluded if they had moderate or severe mitral-valve disease, a creatinine level, >2.3 mg per deciliter [200 μmol per liter] or contraindications to statin therapy. All the participants provided written informed consent before enrollment. The trial protocol was approved by the ethics committees at Fuwai Hospital in Beijing (where patients were recruited) and at the University of Oxford in the United Kingdom (where all electrocardiographic, blood, and statistical analyses were performed).

Eligible patients underwent transthoracic echocardiography for evaluation of left ventricular ejection fraction and left atrial size. Any prescribed statin therapy was stopped, and patients were then randomly assigned to receive rosuvastatin at a dose of 20 mg once daily or matching placebo tablets for up to 8 days before surgery and for 5 days thereafter. Blood samples were obtained at randomization and at 6, 24, 48, and 120 hours after surgery.

### Biomarker analysis

Cystatin C (an alternative biomarker to creatinine for diagnosis of AKI, with some advantages in the estimation of GFR^29^ particularly in the context of an acute physiological insult^30^), growth differentiation factor 15 (GDF-15; a member of the TGF-β cytokine superfamily that may be induced in response to tissue injury and is associated with chronic kidney disease progression^31^), interleukin-6 (IL-6; a pleiotropic cytokine with local and systemic expression that is linked to renal ischemia and AKI^32^), procalcitonin (PCT; a marker of systemic inflammatory response^33^), placental growth factor (PLGF; a proatherogenic cytokine which is present at higher concentrations in individuals with kidney disease^34^), kidney injury molecule-1 (KIM-1; a blood biomarker that specifically reflects renal proximal tubular injury^35^ and is associated with progression to renal failure^36^), and neutrophil gelatinase-associated lipocalin (NGAL; a biomarker of renal epithelial injury^37^) were measured in 960 patients allocated to rosuvastatin and 962 patients allocated to placebo at baseline and after surgery. Other biochemical markers investigated included total creatine kinase (CK; which is linked to an increased risk of AKI in the context of rhabdomyolysis^38^), N-terminal pro-BNP (NT-proBNP), creatinine, LDL cholesterol, and Troponin I. Follow-up values were at 48 hours for CK, cystatin C, KIM-1, and NGAL, and at 6 hours for GDF-15, IL-6, PLGF and PCT.

All blood assays were carried out by the CTSU’s Wolfson Laboratories, University of Oxford, blind to the study treatment allocation. Serum and plasma were separated by centrifugation at 1300g for 10 minutes at room temperature. Further details of the assays used and their respective uncertainties are contained in the **Supplementary Methods**.

### Definition of acute kidney injury (AKI)

AKI by serum creatinine was defined based on KDIGO criteria^39^. Creatinine-defined AKI at 48 hours was defined as an increase from baseline in the creatinine level of ≥0.3 mg per deciliter (≥26.5 µmol per liter) or an increase by a factor of at least 1.5. Creatinine-defined AKI severity was sub-divided based on increase from baseline creatinine by a factor of less than 2 (stage 1); increase from baseline creatinine by a factor of 2 to 3 (stage 2); or increase from baseline creatinine by a factor of more than 3, a rise to a creatinine level of ≥4.0 mg per deciliter (≥350 µmol per liter), or the initiation of renal-replacement therapy (stage 3).

AKI was also separately defined using serum cystatin C^40^. In this case, AKI at 48 hours was defined as an increase from baseline in the cystatin C level by a factor of at least 1.5. Cystatin C-defined AKI severity was sub-divided based on increase from baseline cystatin C by a factor of less than 2 (stage 1); increase from baseline cystatin C by a factor of 2 to 3 (stage 2); or increase from baseline cystatin C by a factor of more than 3, or the initiation of renal-replacement therapy (stage 3). A sensitivity analysis with an alternative definition of stage 1 AKI, to include an increase from baseline in the Cystatin C level by a factor of at least 1.1 to less than 2, was also undertaken.

### Statistical analysis

All randomized comparisons were performed according to the intention-to-treat principle. We used odds ratios and 95% confidence intervals for between-group comparisons of post-operative AKI. ANCOVA was used to compare biomarkers after surgery, with adjustment for baseline values. Analyses were performed on the log scale for all biomarkers and then transformed back to the original scale as geometric means. For dichotomous outcomes, patients with missing data were assumed not to have had the outcome. Missing values for biomarkers were estimated by means of multiple imputation, with 10 replicate sets and combination across sets with the use of Rubin’s methods^41^.

Comparisons of post-operative AKI, creatinine, and cystatin C between treatment groups were performed in subgroups defined according to age at baseline (≤60 years vs >60 years), sex, previous statin use (yes vs no), troponin I concentration (≤0.04 ng per milliliter vs >0.04 ng per milliliter), duration of preoperative randomly assigned regimen (≤2 days vs >2 days), type of surgery (on-pump vs off-pump; CABG vs. aortic-valve replacement), postoperative use of nonsteroidal anti-inflammatory drugs (NSAIDs) or glucocorticoids (yes vs no), and AKI risk in those who received CABG and/or AVR (low/medium/high). Observed effects in different subgroups were compared with the use of chi-square tests for heterogeneity or trend.

To identify factors associated with post-operative AKI risk, analyses were restricted to 1,852 patients who underwent CABG or AVR surgery (or both). Patients with missing creatinine or cystatin C at baseline and/or 48 hours were assumed not to have had the outcome (see **Supplementary Table 1** for numbers of participants with missing values). For dichotomous variables (medical history and medication use) missing values were replaced with the mode. Missing biomarkers were replaced with the mean/median. Separate age-adjusted logistic regression models were used to identify which baseline variables were associated with odds of AKI. Stepwise multivariable forwards and backwards selection models including only those variables found to be significantly associated with odds of AKI at the 5% level in age-adjusted models were then used to develop the final prediction model (selecting the model with the lowest Bayesian Information Criterion). Age was retained in the multivariable model irrespective of its statistical significance. Model discrimination for the final prediction model was assessed using the C statistic. Using the final prediction model, 3 risk groups (low, medium and high) were defined using tertiles of predicted risk.

Cross tabulations of the extent of increase in CK at 48 hours with AKI stage were used to assess the association between CK elevations and post-operative AKI. The final model was additionally adjusted for increases in CK to estimate the relevance of treatment allocation to odds of AKI independent of CK elevations.

## Results

Baseline characteristics of the 1922 patients who underwent randomization are shown in **Table 1**. After randomization, surgery was cancelled for 48 patients (28 patients in the rosuvastatin group and 20 in the placebo group). Of those who received surgery, 1614 (86%) underwent CABG (including 53 [3%] who, in addition, had AVR), 185 (10%) underwent AVR alone, and a further 75 (4%) underwent some other type of surgery; 55% of the operations were on-pump. Overall 66% of patients were statin-naïve (639/960 in the rosuvastatin group and 630/962 in the placebo group). The prevalence of self-reported chronic kidney disease (CKD) was ≤1% in both groups (10/960 in the rosuvastatin group and 8/962 in the placebo group). The number of participants in whom each biomarker result was missing at baseline and follow-up in each group are reported in **Supplementary Table 1**; overall, >91% of results were available.

**Table 1:** Characteristics of the patients at baseline.

| **Characteristic** | **Rosuvastatin**<br>**(n=960)** | **Placebo**<br>**(n=962)** |
| --- | --- | --- |
| Age |  |  |
| Mean – years | 59.3±9.4 | 59.5±9.5 |
| ≤60 years | 516 (53.8%) | 504 (52.4%) |
| >60 years | 444 (46.3%) | 458 (47.6%) |
| Female sex | 194 (20.2%) | 205 (21.3%) |
| Body-mass index, kg/m<sup>2</sup> | 25.7±3.2 | 25.7±3.1 |
| CKD-EPI estimated GFR (ml/min/1.73m<sup>2</sup>)‡ | 90±15 | 90±15 |
| Current smoker | 229 (23.9%) | 245 (25.5%) |
| Medical history |  |  |
| Hypertension | 621 (64.7%) | 614 (63.8%) |
| Myocardial infarction | 282 (29.4%) | 274 (28.5%) |
| Stroke/transient ischaemic attacks | 115 (12.0%) | 119 (12.4%) |
| Peripheral arterial disease | 23 (2.4%) | 19 (2.0%) |
| Heart failure | 46 (4.8%) | 37 (3.8%) |
| Chronic obstructive pulmonary disease | 5 (0.5%) | 14 (1.5%) |
| Diabetes mellitus | 310 (32.3%) | 291 (30.2%) |
| Chronic kidney disease | 10 (1.0%) | 8 (0.8%) |
| Contrast agents (last 2 weeks) | 383 (39.9%) | 379 (39.4%) |
| Current/recent medication§ |  |  |
| Beta-blocker | 813 (84.7%) | 804 (83.6%) |
| NSAID or glucocorticoid | 16 (1.7%) | 7 (0.7%) |
| Insulin | 140 (14.6%) | 158 (16.4%) |
| Antiplatelet or anticoagulant | 792 (82.5%) | 779 (81.0%) |
| Calcium-channel blocker | 445 (46.4%) | 424 (44.1%) |
| ACE inhibitor or ARB | 385 (40.1%) | 384 (39.9%) |
| Nitrate | 798 (83.1%) | 800 (83.2%) |
| Diuretic agent | 197 (20.5%) | 213 (22.1%) |
| Statin | 321 (33.4%) | 332 (34.5%) |
| Scheduled surgery |  |  |
| On-pump procedure | 508 (52.9%) | 515 (53.5%) |
| Off-pump procedure | 422 (44.0%) | 429 (44.6%) |
| CABG | 832 (86.7%) | 838 (87.1%) |
| Aortic-valve replacement | 117 (12.2%) | 119 (12.4%) |

Plus-minus values are means ±SD.
‡ The glomerular filtration rate (GFR) was estimated with the Chronic Kidney Disease Epidemiology Collaboration (CKD-EPI) equation.
§ Recent medication included antiplatelet agents, which are routinely stopped 5 to 7 days before surgery.
ACE denotes angiotensin converting enzyme, ARB angiotensin-receptor blocker, CABG coronary-artery bypass grafting, and NSAID nonsteroidal anti-inflammatory drug.
Table reproduced with permission from Zheng et al.<sup>17</sup>

### Post-operative renal function and incidence of AKI

The incidence of creatinine-defined AKI was greater in the rosuvastatin group than in the placebo group (24.7% vs 19.3%, odds ratio [OR] 1.37, 95% confidence interval [CI] 1.10-1.70, p=0.005; **Figure 1A**), as was the incidence of cystatin C defined AKI (9.2% vs 5.1%, OR 1.86, 95% CI 1.29-2.67, p<0.001; **Figure 1B**). In a sensitivity analysis in which the cystatin C definition of AKI was expanded to include an increase from baseline in the cystatin C level by a factor of at least 1.1, the incidence of AKI was 46.0% vs 36.7% (OR 1.47, 95% CI 1.23-1.77, p<0.001). Creatinine and cystatin C levels in both treatment groups increased from baseline to 48 hours after surgery; however, baseline-adjusted creatinine and cystatin C levels at 48 hours were significantly higher in rosuvastatin-allocated than placebo-allocated patients (1.02±0.01 mg/dL vs 0.99±0.01 mg/dL, p=0.007, **Figure 1C**; and 1.07±0.01 mg/L vs 1.02±0.01 mg/L, p<0.001, **Figure 1D**; respectively).

**Figure 1:**
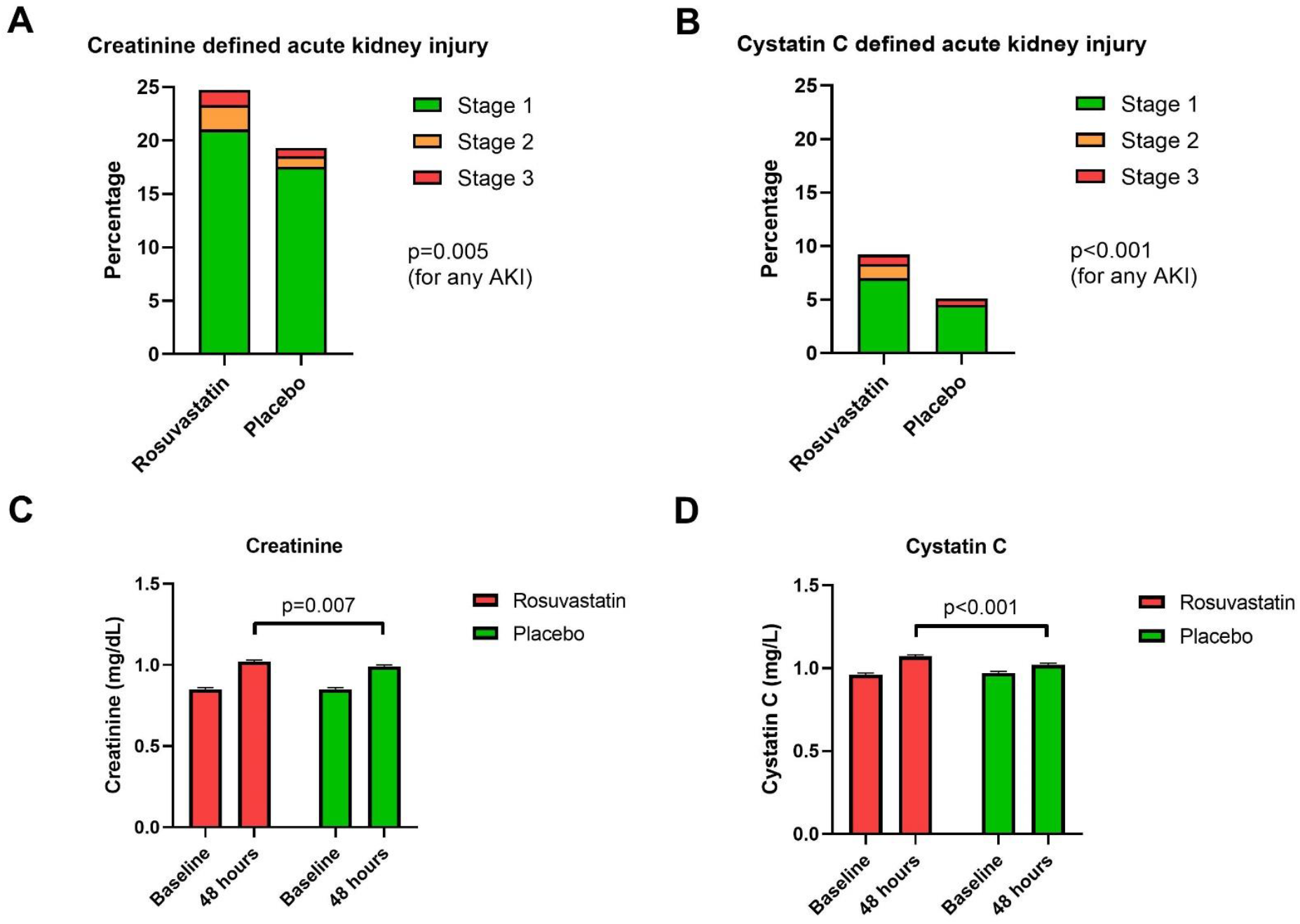
Effect of allocation to rosuvastatin on creatinine-defined and cystatin C-defined AKI, and on post-operative creatinine and cystatin C levels. The upper panel shows bar graphs indicating proportion of AKI stages 1-3 at 48 hours post-operatively in the rosuvastatin and placebo groups, defined by creatinine (**panel A**) and cystatin C (**panel B**). Participants missing cystatin C or creatinine were assumed not to have AKI unless they had undergone renal replacement therapy. The lower panel shows bar graphs indicating levels of creatinine (**panel C**) and cystatin C (**panel D**) at baseline and at 48 hours post-operatively in the rosuvastatin and placebo groups. Bars show geometric means with approximate ±SE. P values were derived from analysis of covariance with adjustment for the baseline value with any missing data imputed with the use of multiple imputation.

The breakdown of absolute excess in creatinine-defined AKI in the rosuvastatin group compared with the placebo group was: stage 1 (3.5±1.8%); stage 2 (1.3±0.6%); and stage 3 (0.5±0.5%). When assessed by cystatin C, the corresponding excess values were: stage 1 (2.5±1.1%); stage 2 (1.3±0.4%); and stage 3 (0.3±0.4%). The proportional effect of allocation to rosuvastatin on post-operative creatinine-defined or cystatin C defined AKI was similar in different types of patients (**Figure 2** and **Supplementary Figure 1**), including in those who were already receiving statins compared to those who were statin-naïve. The effect of allocation to rosuvastatin on mean levels of creatinine and cystatin-C at 48 hours after surgery was also similar in different types of patient (**Supplementary Figure 2** and **Supplementary Figure 3**).

**Figure 2:**
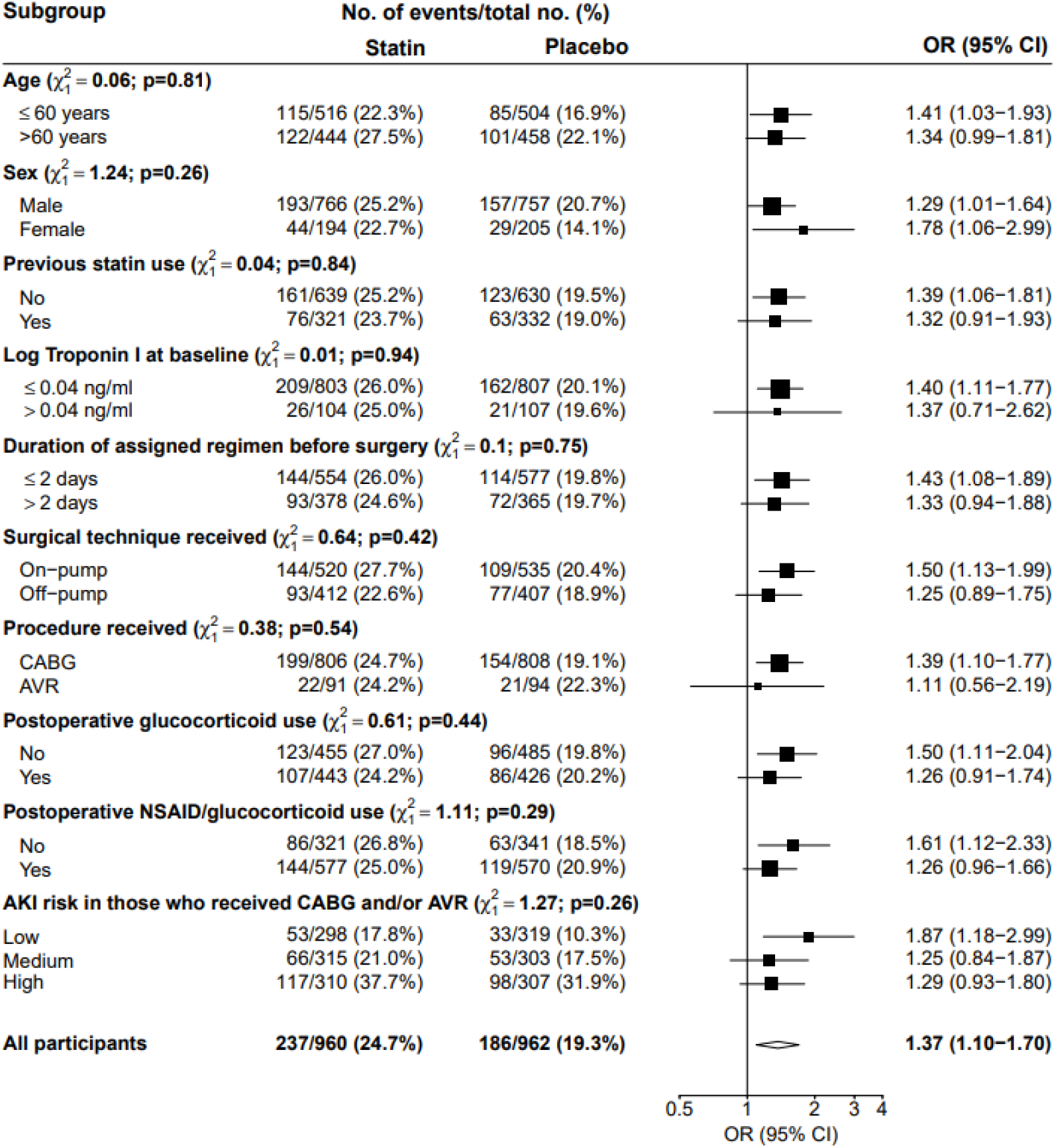
Effect of allocation to rosuvastatin on postoperative creatinine-defined AKI, overall and according to subgroups. Forest plot indicating the effect of rosuvastatin versus placebo on post-operative AKI according to pre-specified subgroups. 142 participants with missing baseline and/or 48hr creatinine levels and who did not undergo renal replacement therapy were assumed to not have had the event. The denominator for coronary-artery bypass grafting (CABG) includes only patients who had CABG without aortic-valve replacement, and the denominator for aortic-valve replacement includes only those who had aortic-valve replacement without CABG. Participants with a missing value for a particular subgroup are not shown (but do contribute to the overall diamond). 101 participants were missing baseline Troponin. 48 participants were scheduled for surgery but did not undergo surgery; these individuals are excluded from the relevant subgroups. Risk groups were defined using the predicted values from a logistic regression that included age and five variables found to be associated with AKI risk in 1,852 participants who underwent CABG or AVR surgery in an age-adjusted multivariate model that included treatment allocation: baseline creatinine, baseline KIM-1, procedure (CABG only/AVR only/both), insulin use, and contrast agent use. Abbreviations: OR, odds ratio; CI, confidence interval.

### Serum biomarkers relevant to post-operative AKI by statin allocation

Concentrations of GDF-15, IL-6, PCT, PLGF, KIM-1 and NGAL were all substantially higher after surgery than at baseline (**Figure 3**). For GDF-15, IL-6, PCT, PLGF and NGAL there was no significant difference in this rise between rosuvastatin-allocated and placebo-allocated patients, but for KIM-1 the rise was significantly higher for patients allocated rosuvastatin (baseline-adjusted mean KIM-1 concentration at 48 hours: 278±5 pg/ml versus 259±5 pg/ml respectively, p=0.01; **Figure 3E**).

**Figure 3:**
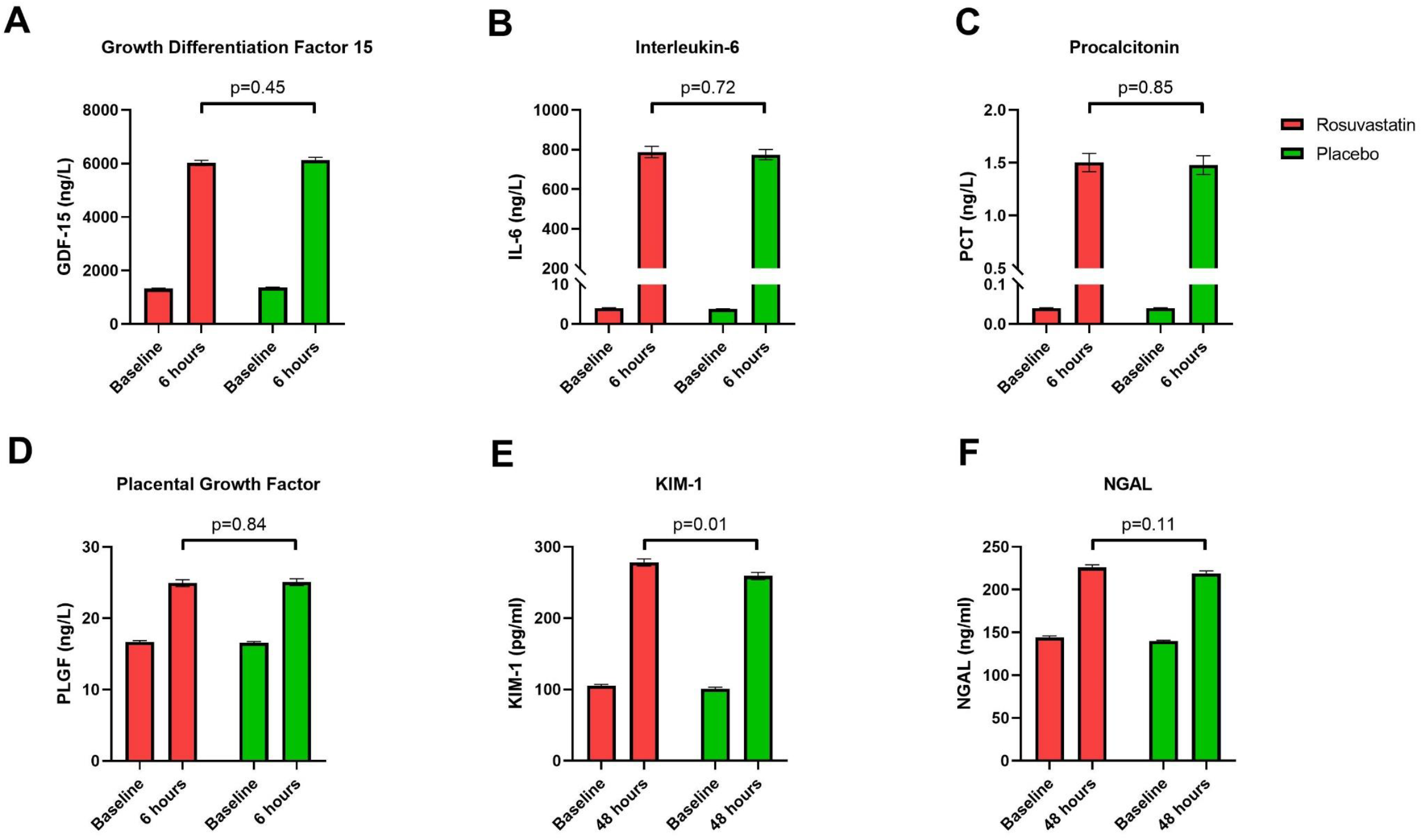
Effect of allocation to rosuvastatin on postoperative concentrations of biomarkers of inflammation and renal injury. Bar graphs indicating levels of growth differentiation factor 15 (GDF-15; **panel A**), interleukin-6 (IL-6; **panel B**), procalcitonin (**panel C**), placental growth factor (PLGF; **panel D**), kidney injury molecule-1 (KIM-1; **panel E**), and neutrophil gelatinase-associated lipocalin (NGAL; **panel F**) at baseline and post-operatively in the rosuvastatin and placebo groups. P values were derived from analysis of covariance with adjustment for the baseline value with any missing data imputed with the use of multiple imputation. Bars show geometric means with approximate ±SE.

The incidence of elevation in CK at 48 hours after surgery to >5x the baseline level was similar in both groups (37.1% in the rosuvastatin group vs 37.5% in the placebo group, p=0.84). However, elevations to >10x and >40x the baseline level were more common in the rosuvastatin group (30.9% versus 26.5%, p=0.032, and 2.1% versus 0.7%, p=0.016, respectively). The extent of increase in CK at 48 hours after surgery was correlated with the likelihood of AKI, both for creatinine-defined AKI (**Supplementary Table 2**) and cystatin C-defined AKI (**Supplementary Table 3**).

### Predictors of post-operative AKI

The individual and joint relevance of clinical factors and baseline biomarkers to the odds of post-operative creatinine and cystatin C-defined AKI were investigated (**Table 2** and **Table 3**, respectively).

**Table 2:**
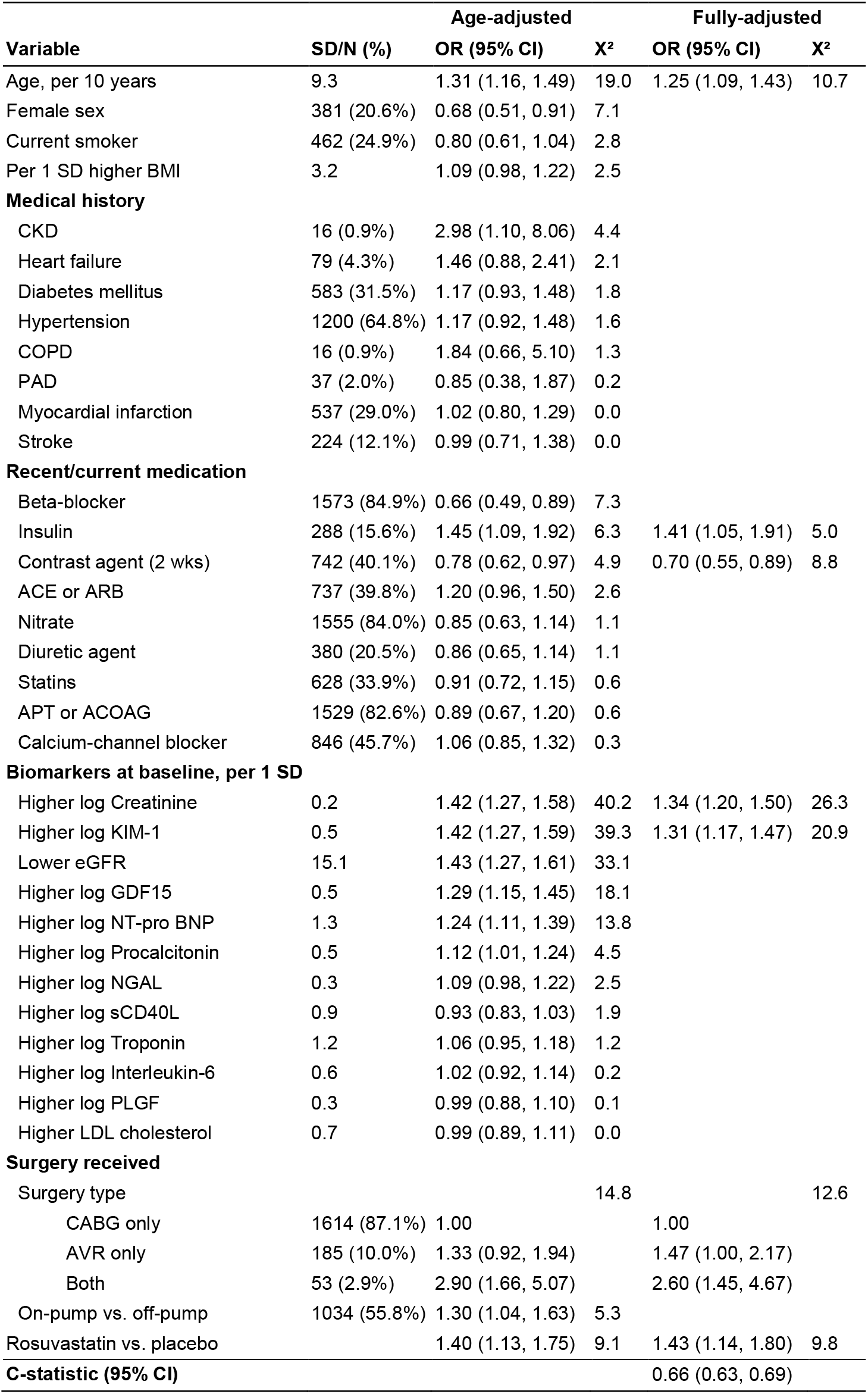
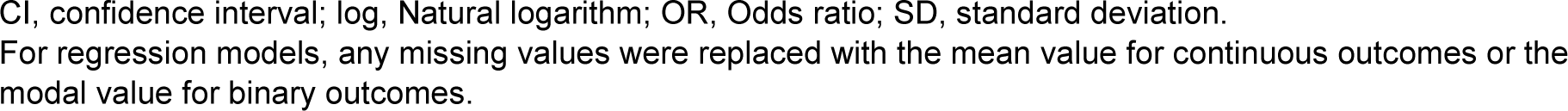
Age- and fully-adjusted relevance of each baseline measure to the odds of creatinine-defined AKI among 1852 participants who had either CABG or AVR (or both)

| Variable | SD/N (%) | Age-adjusted |  | Fully-adjusted |  |
| --- | --- | --- | --- | --- | --- |
|  |  | OR (95% CI) | X<sup>2</sup> | OR (95% CI) | X<sup>2</sup> |
| Age, per 10 years | 9.3 | 1.31 (1.16, 1.49) | 19.0 | 1.25 (1.09, 1.43) | 10.7 |
| Female sex | 381 (20.6%) | 0.68 (0.51, 0.91) | 7.1 |  |  |
| Current smoker | 462 (24.9%) | 0.80 (0.61, 1.04) | 2.8 |  |  |
| Per 1 SD higher BMI | 3.2 | 1.09 (0.98, 1.22) | 2.5 |  |  |
| **Medical history** |  |  |  |  |  |
| CKD | 16 (0.9%) | 2.98 (1.10, 8.06) | 4.4 |  |  |
| Heart failure | 79 (4.3%) | 1.46 (0.88, 2.41) | 2.1 |  |  |
| Diabetes mellitus | 583 (31.5%) | 1.17 (0.93, 1.48) | 1.8 |  |  |
| Hypertension | 1200 (64.8%) | 1.17 (0.92, 1.48) | 1.6 |  |  |
| COPD | 16 (0.9%) | 1.84 (0.66, 5.10) | 1.3 |  |  |
| PAD | 37 (2.0%) | 0.85 (0.38, 1.87) | 0.2 |  |  |
| Myocardial infarction | 537 (29.0%) | 1.02 (0.80, 1.29) | 0.0 |  |  |
| Stroke | 224 (12.1%) | 0.99 (0.71, 1.38) | 0.0 |  |  |
| **Recent/current medication** |  |  |  |  |  |
| Beta-blocker | 1573 (84.9%) | 0.66 (0.49, 0.89) | 7.3 |  |  |
| Insulin | 288 (15.6%) | 1.45 (1.09, 1.92) | 6.3 | 1.41 (1.05, 1.91) | 5.0 |
| Contrast agent (2 wks) | 742 (40.1%) | 0.78 (0.62, 0.97) | 4.9 | 0.70 (0.55, 0.89) | 8.8 |
| ACE or ARB | 737 (39.8%) | 1.20 (0.96, 1.50) | 2.6 |  |  |
| Nitrate | 1555 (84.0%) | 0.85 (0.63, 1.14) | 1.1 |  |  |
| Diuretic agent | 380 (20.5%) | 0.86 (0.65, 1.14) | 1.1 |  |  |
| Statins | 628 (33.9%) | 0.91 (0.72, 1.15) | 0.6 |  |  |
| APT or ACOAG | 1529 (82.6%) | 0.89 (0.67, 1.20) | 0.6 |  |  |
| Calcium-channel blocker | 846 (45.7%) | 1.06 (0.85, 1.32) | 0.3 |  |  |
| **Biomarkers at baseline, per 1 SD** |  |  |  |  |  |
| Higher log Creatinine | 0.2 | 1.42 (1.27, 1.58) | 40.2 | 1.34 (1.20, 1.50) | 26.3 |
| Higher log KIM-1 | 0.5 | 1.42 (1.27, 1.59) | 39.3 | 1.31 (1.17, 1.47) | 20.9 |
| Lower eGFR | 15.1 | 1.43 (1.27, 1.61) | 33.1 |  |  |
| Higher log GDF15 | 0.5 | 1.29 (1.15, 1.45) | 18.1 |  |  |
| Higher log NT-pro BNP | 1.3 | 1.24 (1.11, 1.39) | 13.8 |  |  |
| Higher log Procalcitonin | 0.5 | 1.12 (1.01, 1.24) | 4.5 |  |  |
| Higher log NGAL | 0.3 | 1.09 (0.98, 1.22) | 2.5 |  |  |
| Higher log sCD40L | 0.9 | 0.93 (0.83, 1.03) | 1.9 |  |  |
| Higher log Troponin | 1.2 | 1.06 (0.95, 1.18) | 1.2 |  |  |
| Higher log Interleukin-6 | 0.6 | 1.02 (0.92, 1.14) | 0.2 |  |  |
| Higher log PLGF | 0.3 | 0.99 (0.88, 1.10) | 0.1 |  |  |
| Higher LDL cholesterol | 0.7 | 0.99 (0.89, 1.11) | 0.0 |  |  |
| **Surgery received** |  |  |  |  |  |
| Surgery type |  |  | 14.8 |  | 12.6 |
| CABG only | 1614 (87.1%) | 1.00 |  | 1.00 |  |
| AVR only | 185 (10.0%) | 1.33 (0.92, 1.94) |  | 1.47 (1.00, 2.17) |  |
| Both | 53 (2.9%) | 2.90 (1.66, 5.07) |  | 2.60 (1.45, 4.67) |  |
| On-pump vs. off-pump | 1034 (55.8%) | 1.30 (1.04, 1.63) | 5.3 |  |  |
| Rosuvastatin vs. placebo |  | 1.40 (1.13, 1.75) | 9.1 | 1.43 (1.14, 1.80) | 9.8 |
| **C-statistic (95% CI)** |  |  |  | 0.66 (0.63, 0.69) |  |

CI, confidence interval; log, Natural logarithm; OR, Odds ratio; SD, standard deviation. For regression models, any missing values were replaced with the mean value for continuous outcomes or the modal value for binary outcomes.

**Table 3:**
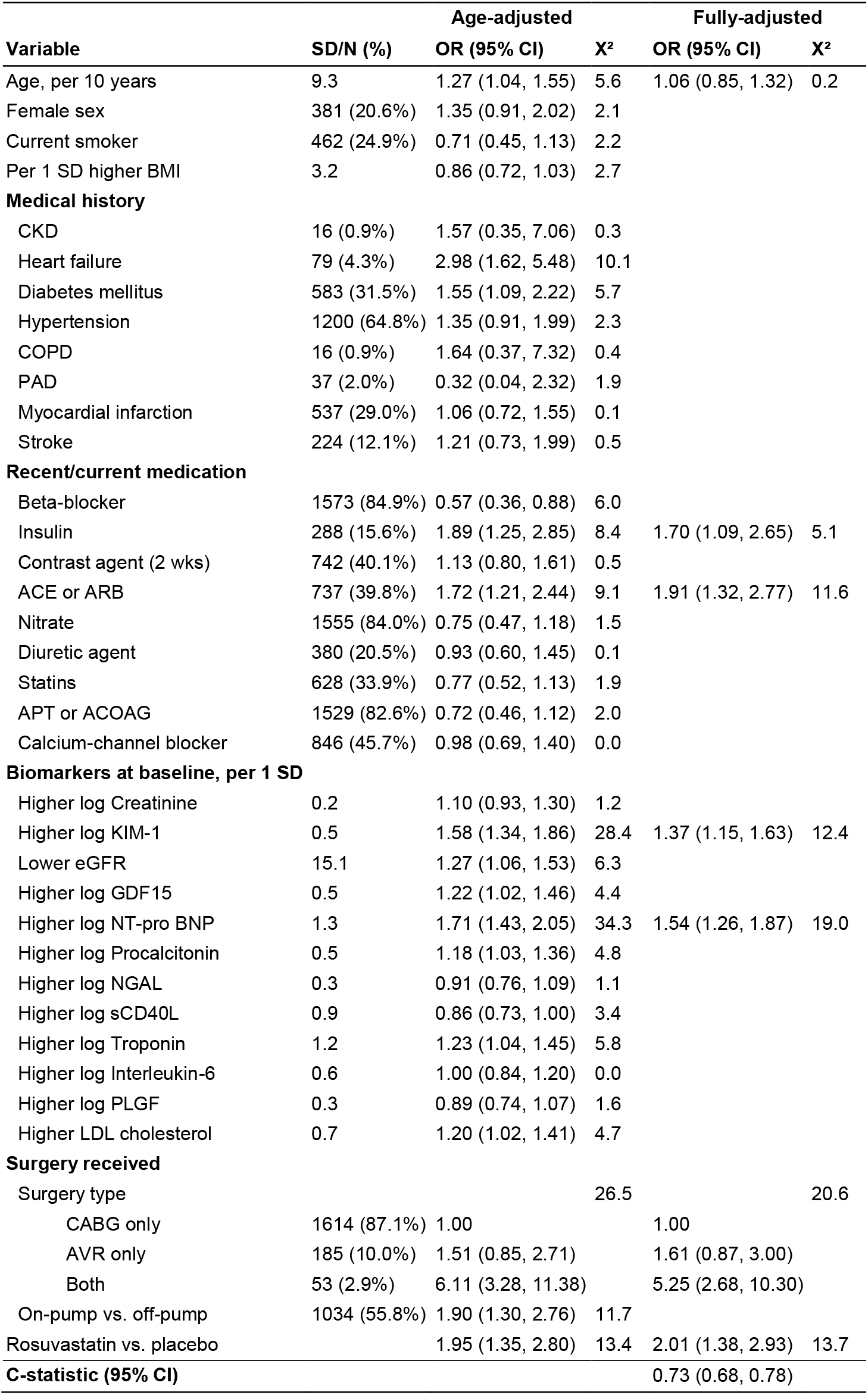
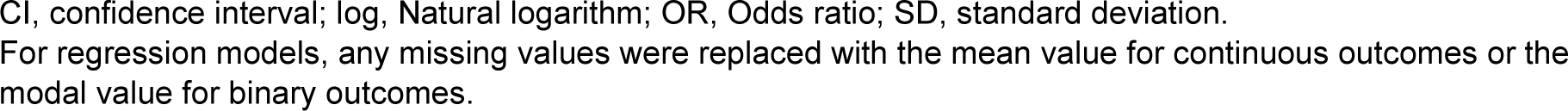
Age- and fully-adjusted relevance of each baseline measure to the odds of cystatin C-defined AKI among 1852 participants who had either CABG or AVR (or both)

| Variable | SD/N (%) | Age-adjusted |  | Fully-adjusted |  |
| --- | --- | --- | --- | --- | --- |
|  |  | OR (95% CI) | X<sup>2</sup> | OR (95% CI) | X<sup>2</sup> |
| Age, per 10 years | 9.3 | 1.27 (1.04, 1.55) | 5.6 | 1.06 (0.85, 1.32) | 0.2 |
| Female sex | 381 (20.6%) | 1.35 (0.91, 2.02) | 2.1 |  |  |
| Current smoker | 462 (24.9%) | 0.71 (0.45, 1.13) | 2.2 |  |  |
| Per 1 SD higher BMI | 3.2 | 0.86 (0.72, 1.03) | 2.7 |  |  |
| **Medical history** |  |  |  |  |  |
| CKD | 16 (0.9%) | 1.57 (0.35, 7.06) | 0.3 |  |  |
| Heart failure | 79 (4.3%) | 2.98 (1.62, 5.48) | 10.1 |  |  |
| Diabetes mellitus | 583 (31.5%) | 1.55 (1.09, 2.22) | 5.7 |  |  |
| Hypertension | 1200 (64.8%) | 1.35 (0.91, 1.99) | 2.3 |  |  |
| COPD | 16 (0.9%) | 1.64 (0.37, 7.32) | 0.4 |  |  |
| PAD | 37 (2.0%) | 0.32 (0.04, 2.32) | 1.9 |  |  |
| Myocardial infarction | 537 (29.0%) | 1.06 (0.72, 1.55) | 0.1 |  |  |
| Stroke | 224 (12.1%) | 1.21 (0.73, 1.99) | 0.5 |  |  |
| **Recent/current medication** |  |  |  |  |  |
| Beta-blocker | 1573 (84.9%) | 0.57 (0.36, 0.88) | 6.0 |  |  |
| Insulin | 288 (15.6%) | 1.89 (1.25, 2.85) | 8.4 | 1.70 (1.09, 2.65) | 5.1 |
| Contrast agent (2 wks) | 742 (40.1%) | 1.13 (0.80, 1.61) | 0.5 |  |  |
| ACE or ARB | 737 (39.8%) | 1.72 (1.21, 2.44) | 9.1 | 1.91 (1.32, 2.77) | 11.6 |
| Nitrate | 1555 (84.0%) | 0.75 (0.47, 1.18) | 1.5 |  |  |
| Diuretic agent | 380 (20.5%) | 0.93 (0.60, 1.45) | 0.1 |  |  |
| Statins | 628 (33.9%) | 0.77 (0.52, 1.13) | 1.9 |  |  |
| APT or ACOAG | 1529 (82.6%) | 0.72 (0.46, 1.12) | 2.0 |  |  |
| Calcium-channel blocker | 846 (45.7%) | 0.98 (0.69, 1.40) | 0.0 |  |  |
| **Biomarkers at baseline, per 1 SD** |  |  |  |  |  |
| Higher log Creatinine | 0.2 | 1.10 (0.93, 1.30) | 1.2 |  |  |
| Higher log KIM-1 | 0.5 | 1.58 (1.34, 1.86) | 28.4 | 1.37 (1.15, 1.63) | 12.4 |
| Lower eGFR | 15.1 | 1.27 (1.06, 1.53) | 6.3 |  |  |
| Higher log GDF15 | 0.5 | 1.22 (1.02, 1.46) | 4.4 |  |  |
| Higher log NT-pro BNP | 1.3 | 1.71 (1.43, 2.05) | 34.3 | 1.54 (1.26, 1.87) | 19.0 |
| Higher log Procalcitonin | 0.5 | 1.18 (1.03, 1.36) | 4.8 |  |  |
| Higher log NGAL | 0.3 | 0.91 (0.76, 1.09) | 1.1 |  |  |
| Higher log sCD40L | 0.9 | 0.86 (0.73, 1.00) | 3.4 |  |  |
| Higher log Troponin | 1.2 | 1.23 (1.04, 1.45) | 5.8 |  |  |
| Higher log Interleukin-6 | 0.6 | 1.00 (0.84, 1.20) | 0.0 |  |  |
| Higher log PLGF | 0.3 | 0.89 (0.74, 1.07) | 1.6 |  |  |
| Higher LDL cholesterol | 0.7 | 1.20 (1.02, 1.41) | 4.7 |  |  |
| **Surgery received** |  |  |  |  |  |
| Surgery type |  |  | 26.5 |  | 20.6 |
| CABG only | 1614 (87.1%) | 1.00 |  | 1.00 |  |
| AVR only | 185 (10.0%) | 1.51 (0.85, 2.71) |  | 1.61 (0.87, 3.00) |  |
| Both | 53 (2.9%) | 6.11 (3.28, 11.38) |  | 5.25 (2.68, 10.30) |  |
| On-pump vs. off-pump | 1034 (55.8%) | 1.90 (1.30, 2.76) | 11.7 |  |  |
| Rosuvastatin vs. placebo |  | 1.95 (1.35, 2.80) | 13.4 | 2.01 (1.38, 2.93) | 13.7 |
| **C-statistic (95% CI)** |  |  |  | 0.73 (0.68, 0.78) |  |

CI, confidence interval; log, Natural logarithm; OR, Odds ratio; SD, standard deviation. For regression models, any missing values were replaced with the mean value for continuous outcomes or the modal value for binary outcomes.

In the fully-adjusted multivariable analysis, age, insulin treatment, lack of recent exposure to contrast agent, log baseline creatinine, log baseline KIM-1, combined CABG and AVR surgery, and allocation to rosuvastatin were all independently associated with significantly higher odds of creatinine-defined AKI. The overall C-statistic corresponding to this final model was 0.66 (95% CI 0.63-0.69). In this fully-adjusted model, the conditional odds ratio associated with allocation to rosuvastatin was 1.43 (95% CI 1.14 – 1.80). This was not materially altered by further adjustment for increases in CK (OR with adjustment for CK increase 1.34 [95% CI 1.07-1.69]).

When considering cystatin C-defined AKI, insulin treatment, ACE or ARB treatment, log baseline KIM-1, log baseline NT-pro BNP, combined CABG and AVR surgery, and allocation to rosuvastatin were all independently associated with significantly higher odds of AKI. The overall C-statistic corresponding to this final model was 0.73 (95% CI 0.68-0.78). In this fully-adjusted model, the conditional odds ratio associated with allocation to rosuvastatin was 2.01 (95% CI 1.38 – 2.93). This was not materially altered by further adjustment for increases in CK (OR with adjustment for CK increase 1.92 [95% CI 1.31-2.81]).

Overall, insulin treatment, baseline KIM-1, combined CABG and AVR surgery, and allocation to rosuvastatin were all independently associated with AKI by any definition (**Table 4**).

**Table 4:** Fully-adjusted relevance of selected baseline measures to the odds of creatinine-defined and cystatin C-defined AKI among 1852 participants who had either CABG or AVR (or both)

| Variable | SD/N (%) | Creatinine-defined AKI |  | Cystatin C-defined AKI |  |
| --- | --- | --- | --- | --- | --- |
|  |  | OR (95% CI) | X<sup>2</sup> | OR (95% CI) | X<sup>2</sup> |
| Age, per 10 years | 9.3 | 1.25 (1.09, 1.43) | 10.7 |  |  |
| **Insulin** | 288 (15.6%) | **1.41 (1.05, 1.91)** | **5.0** | **1.70 (1.09, 2.65)** | **5.1** |
| Contrast agent (2 wks) | 742 (40.1%) | 0.70 (0.55, 0.89) | 8.8 |  |  |
| ACE or ARB | 737 (39.8%) |  |  | 1.91 (1.32, 2.77) | 11.6 |
| Higher log Creatinine | 0.2 | 1.34 (1.20, 1.50) | 26.3 |  |  |
| **Higher log KIM-1** | 0.5 | **1.31 (1.17, 1.47)** | **20.9** | **1.37 (1.15, 1.63)** | **12.4** |
| Higher log NT-pro BNP | 1.3 |  |  | 1.54 (1.26, 1.87) | 19.0 |
| **Surgery type** |  |  | **12.6** |  | **20.6** |
| CABG only | 1614 (87.1%) | 1.00 |  | 1.00 |  |
| AVR only | 185 (10.0%) | 1.47 (1.00, 2.17) |  | 1.61 (0.87, 3.00) |  |
| **Both** | 53 (2.9%) | **2.60 (1.45, 4.67)** |  | **5.25 (2.68, 10.30)** |  |
| **Rosuvastatin vs. placebo** |  | **1.43 (1.14, 1.80)** | **9.8** | **2.01 (1.38, 2.93)** | **13.7** |

CI, confidence interval; log, Natural logarithm; OR, Odds ratio; SD, standard deviation.
For regression models, any missing values were replaced with the mean value for continuous outcomes or the modal value for binary outcomes. Variables were selected for inclusion in this Table if they were statistically significant in either the fully-adjusted model for creatinine-defined AKI or in the fully-adjusted model for cystatin C-defined AKI; variables that were statistically significant in both models are highlighted in bold.

When risk factors were combined to create risk scores for AKI and participants were divided into tertiles of predicted risk based on these scores, the effect of allocation to rosuvastatin compared to placebo was similar across risk groups (p for trend = 0.26 for creatinine-defined AKI, **Figure 2**; p for trend = 0.13 for cystatin C-defined AKI, **Supplementary Figure 1**).

## Discussion

This study demonstrated that allocation to rosuvastatin compared with placebo increased the absolute risk of post-operative AKI by 4-5% in patients undergoing cardiac surgery, and led to higher post-operative KIM-1, suggesting a deleterious effect of rosuvastatin on renal function possibly mediated by proximal tubular injury^36^. In multivariable analyses, insulin treatment, baseline KIM-1, combined CABG and AVR surgery, and allocation to rosuvastatin were all independently associated with AKI by any definition. Although rosuvastatin also led to a significant increase in post-operative CK, adjustment for this did not materially change the estimated effect of allocation to rosuvastatin on post-operative AKI. Overall, the adverse effect of perioperative rosuvastatin on renal function is particularly relevant given its lack of efficacy in terms of reducing postoperative AF or perioperative myocardial injury^17^.

The mechanism of AKI in the postoperative patient is usually multi-factorial, and often related to renal ischaemia, nephrotoxic medications, hypovolaemia, and sepsis^42^. Cardiac surgery and major vascular surgery are particularly associated with risk of AKI, probably due to frequent combination of underlying chronic kidney disease (possibly vascular in origin) and the degree of insult resulting from the surgery. In line with this paradigm, previously identified risk factors for development of AKI include age, congestive heart failure, prior myocardial revascularisation, type 1 diabetes mellitus or pre-operative hyperglycaemia, and elevated pre-operative serum creatinine^43^. Interventions to reduce risk of postoperative AKI are limited and largely conservative, based around avoidance of nephrotoxic medications, hyperglycaemia, and hypo- or hypervolaemia.

In the present study, we measured a variety of additional novel biomarkers in order to further investigate potential mechanisms of AKI in this patient group. In addition to creatinine, serum cystatin C was also used to diagnose AKI, as the latter may be less affected by non-GFR-related factors such as age, gender, muscle mass, and ethnicity^29^. Baseline KIM-1 was independently associated with the risk of post-operative AKI, in keeping with previous data suggesting that KIM-1 is a biomarker of acute and chronic kidney injury^35, 36^. The post-operative concentration of KIM-1 was also higher in patients allocated to rosuvastatin compared to placebo, suggesting that allocation to rosuvastatin may exacerbate renal proximal tubular injury^35^ in the context of cardiac surgery. These results are in contrast to those for NGAL, which is a marker of renal epithelial tissue injury. Similarly, there were also no significant differences in serum GDF-15, IL6, PCT, or PLGF between the groups, suggesting that the adverse effect of perioperative rosuvastatin on renal function may be independent of systemic inflammation and renal epithelial tissue injury.

These findings are contrary to prior interest in the use of statins to reduce post-operative complications in patients undergoing high risk surgery based on the described pleiotropic effects of statins, including anti-inflammatory^5^ and antioxidant^6, 7^ effects. Current ESC and ACC/AHA practice guidelines on the cardiovascular evaluation and management of patients undergoing non-cardiac surgery both recommend consideration of initiation of statin therapy in the peri-operative period in individuals undergoing vascular surgery who are not already receiving long-term statin therapy^44, 45^.

Beyond the context of cardiac surgery, myotoxicity is recognized as a rare side-effect of statin therapy^46^. This is heterogeneous in presentation but can rarely result in rhabdomyolysis, characterized by the leakage of muscle-cell contents (including CK and myoglobin) into the circulation^47^. An increase in serum myoglobin concentration is described after cardiac surgery, and independently predicts the incidence of postoperative AKI^48^. Although allocation to rosuvastatin in STICS did significantly increase post-operative CK, adjustment for this increase in CK did not significantly attenuate the effect of rosuvastatin on risk of postoperative AKI, implying that a statin-associated muscle injury does not contribute to the latter.

The finding that perioperative initiation of rosuvastatin 20mg increases the risk of AKI after cardiac surgery is clinically relevant, particularly in the context of a null effect of statin therapy on post-operative complications^17^. Although previous randomized controlled trials have demonstrated a renal protective effect of statins on contrast-induced AKI^49^, the present data indicate that rosuvastatin has an adverse effect on renal function in patients undergoing cardiac surgery, perhaps reflecting differences in the nature and physiological insult of the procedure itself. A systematic review and meta-analysis of 11 trials of perioperative statin therapy in cardiac surgery (including STICS) confirmed a higher incidence of AKI in cardiac surgery patients receiving perioperative statins compared to control (RR = 1.15 [95% CI 1.00-1.31], p=0.05), with no significant heterogeneity between trials^50^. In addition to their small size, many of the trials had other important limitations including a failure to include AKI as a pre-specified outcome with a clear definition of the diagnostic criteria (7 trials), lack of confirmation of an intention-to-treat analysis (6 trials), and lack of allocation concealment or blinding (5 trials). However, when the meta-analysis was limited to the 5 trials (including STICS) that were assessed as having a low risk of bias, a similar result was seen (RR = 1.17 [95% CI 1.02-1.34], p=0.03)^50^. STICS contributed the strongest evidence on this topic by including 1922 of 3384 (57%) patients and more importantly also accounting for 423 of 650 (65%) AKI events. Overall, the size and robustness of the STICS trial provided a unique opportunity to further investigate the mechanisms and predictors of AKI in the present study.

It is important to emphasise that the majority of the excess in AKI following rosuvastatin use in STICS was relatively minor (i.e. stage 1). Whilst incompletely resolved AKI could lead to CKD and increase the risk of future cardiovascular events, it is likely that the prognostic benefit from statin therapy in terms of long-term LDL reduction would outweigh any such risk. The STICS trial does not directly address the question of whether existing statin therapy should be temporarily suspended at the time of cardiac surgery, since only approximately one third of patients were already prescribed statins, although there was no evidence of a differential effect on AKI in this subgroup compared to the statin-naïve group. In the absence of further trials, temporary statin cessation in the perioperative period in patients undergoing elective cardiac surgery may be a reasonable option to consider on a case-by-case basis, particularly in patients with several other risk factors for development of AKI. If such measures were undertaken, close attention would need to be paid to statin reintroduction after the risk of perioperative AKI has receded.

This study has a number of strengths including its large size, high levels of adherence to the treatment regimen, robust and blinded evaluation of AKI (as a pre-specified outcome) using both creatinine and serum cystatin C, and evaluation of a number of other biomarkers that are potentially mechanistically linked to development of postoperative AKI. Limitations include lack of urine samples to explore other biomarkers of renal tubular function, and a lack of data regarding post-operative urine output, which could therefore not be incorporated within the definitions of AKI used in the study. It is not possible to determine from this dataset whether the findings reported are specific to the patients’ ethnic group and/or to rosuvastatin (versus other statins), and information is also not available on longer-term renal outcomes after the perioperative period.

In summary, perioperative rosuvastatin initiation increased the absolute risk of AKI (however defined) after cardiac surgery by 4-5%, and led to higher post-operative KIM-1, suggesting a deleterious effect on renal function possibly mediated by proximal tubular injury. Insulin treatment, baseline KIM-1, combined CABG and AVR surgery, and allocation to rosuvastatin were all independently associated with AKI by any definition.

## Data Availability

The data underlying this article will be shared on reasonable request to the corresponding author.

## Acknowledgements

We thank the patients who agreed to take part in the trial; the cardiac surgery teams in the Department of Cardiac Surgery, Fuwai Hospital, Beijing; the research support staff in the China–Oxford Center for International Health Research, Fuwai Hospital; and the technicians in the Wolfson Laboratories of the Clinical Trial Service Unit and Epidemiological Studies Unit, Oxford, United Kingdom.

