## Supplementary Material for "Mechanisms and Predictors of Acute Kidney Injury with Perioperative Rosuvastatin in Patients Undergoing Cardiac Surgery"

**Supplemental Material**

**Supplementary Methods**

*Assay details*

Troponin I was measured in serum using a chemiluminescent immunoassay on a Beckman Coulter Access 2 AccuTnI (Reference A78803); the manufacturer's quoted analytical range is 0.01 ng/mL to 100 ng/mL, with a functional sensitivity of 0.03 ng/mL, as determined by a total imprecision of 20% coefficient of variance (which was confirmed in the CTSU laboratories). Plasma was used for measuring LDL cholesterol (direct homogeneous assay) and creatinine (Kinetic Jaffé compensated assay) using a Beckman Coulter AU680. Serum was used for measuring cystatin C (Gentian turbidimetric immunoassay) and CK (kinetic assay) using a Beckman Coulter DxC700AU. PIGF, GDF-15, IL-6, and procalcitonin were measured in serum using electrochemiluminescence immunoassays on a Roche Cobas e601. NT-proBNP was measured in plasma and sCD40L in serum using plate assays with electrochemiluminescence detection (Meso Scale Discovery). Serum was used for measuring NGAL and KIM-1 using Ella Simple Plex sandwich immunoassay cartridges with fluorescent detection (Biotechne). The LDL cholesterol assay used N-geneous reagents, calibrators and settings from Genzyme Diagnostics, UK; the creatinine, Troponin I, cystatin C and CK assays used reagents, calibrators and settings from Beckman Coulter; the PIGF, GDF-15, IL-6, and procalcitonin assays used reagents, calibrators and settings from Roche. At least two levels of quality control material were run for each assay. For each assay, the expanded uncertainty with a coverage factor (k) of 2 is as follows for the levels stated; Troponin I 0.88 ng/mL at 10.1 ng/mL; LDL cholesterol 4.32 mg/dL at 67.94 mg/dL, creatinine 0.06 mg/dL at 1.10 mg/dL, cystatin C 0.22 mg/L at 3.52 mg/L; CK 17.5 U/L at 380 U/L; PIGF 4.8 ng/L at 110.6 ng/L; GDF-15 48 ng/L at 1495 ng/L; IL-6 1.74 ng/L at 38.25 ng/L; procalcitonin 0.049 ng/mL at 0.464 ng/mL; NT-proBNP 128 pg/mL at 292 pg/mL, sCD40L 74.3 pg/mL at 137pg/mL, NGAL 3535.22 pg/mL at 42086 pg/mL; and KIM-1 13.45 pg/mL at 354 pg/mL.

**Supplementary Table 1: Number of participants missing biomarker results**

| | Baseline values | | Follow-up values* | |
|---|---|---|---|---|
| **Biomarker** | **Rosuvastatin (n=960)** | **Placebo (n=962)** | **Rosuvastatin (n=960)** | **Placebo (n=962)** |
| CK | 68 (7.1%) | 62 (6.4%) | 68 (7.1%) | 62 (6.4%) |
| Cystatin C | 68 (7.1%) | 62 (6.4%) | 68 (7.1%) | 62 (6.4%) |
| GDF-15 | 69 (7.2%) | 64 (6.7%) | 81 (8.4%) | 68 (7.1%) |
| IL-6 | 99 (10.3%) | 105 (10.9%) | 111 (11.6%) | 109 (11.3%) |
| KIM-1 | 68 (7.1%) | 62 (6.4%) | 69 (7.2%) | 63 (6.5%) |
| NGAL | 68 (7.1%) | 62 (6.4%) | 69 (7.2%) | 64 (6.7%) |
| PLGF | 69 (7.2%) | 64 (6.7%) | 81 (8.4%) | 68 (7.1%) |
| Procalcitonin | 162 (16.9%) | 172 (17.9%) | 173 (18.0%) | 176 (18.3%) |

* Follow-up values were at 48 hours for Creatine Kinase, Cystatin C, KIM-1, and NGAL, and at 6 hours for Growth Differentiation Factor 15, Interleukin-6, Placenta Growth Factor and Procalcitonin.

### Supplementary Table 2: Increase in CK at 48 hours after surgery by creatinine-defined AKI stage

| Acute kidney injury† | ≤5x baseline | >5x baseline | >10x baseline | >40x baseline | missing | Total |
|---|---|---|---|---|---|---|
| No AKI | 406 (81.9%) | 576 (80.3%) | 379 (68.7%) | 11 (40.7%) | 127 (97.7%) | 1499 (78.0%) |
| Stage 1 | 85 (17.1%) | 128 (17.9%) | 146 (26.4%) | 10 (37.0%) | 1 (0.8%) | 370 (19.3%) |
| Stage 2 | 5 (1.0%) | 11 (1.5%) | 14 (2.5%) | 2 (7.4%) | 0 (0.0%) | 32 (1.7%) |
| Stage 3* | 0 (0.0%) | 2 (0.3%) | 13 (2.4%) | 4 (14.8%) | 2 (1.5%) | 21 (1.1%) |
| Any AKI | 90 (18.1%) | 141 (19.7%) | 173 (31.3%) | 16 (59.3%) | 3 (2.3%) | 423 (22.0%) |
| Total | 496 (100.0%) | 717 (100.0%) | 552 (100.0%) | 27 (100.0%) | 130 (100.0%) | 1922 (100.0%) |

† Acute kidney injury at 48 hours was defined as an increase from baseline in the creatinine level of 0.3 mg or more per deciliter (30 µmol per liter) or an increase by a factor of at least 1.5 to less than 2 (stage 1); an increase from baselinein the creatinine level by a factor of 2 to 3 (stage 2); or an increase from baseline in the creatinine level by a factor of more than 3, a rise to a creatinine level of at least 4.0 mg per deciliter (350 µmol per liter), or the initiation of renal-replacement therapy (stage 3).

*Participants missing creatinine at baseline and/or 48hr were assumed to not have AKI, except 2 participants who had RRT and were classified as stage 3 AKI.

### Supplementary Table 3: Increase in CK at 48 hours after surgery by cystatin C-defined AKI stage

| Acute kidney injury† | ≤5x baseline | >5x baseline | >10x baseline | >40x baseline | missing | Total |
|---|---|---|---|---|---|---|
| No AKI | 473 (95.4%) | 674 (94.0%) | 490 (88.8%) | 20 (74.1%) | 128 (98.5%) | 1785 (92.9%) |
| Stage 1 | 22 (4.4%) | 40 (5.6%) | 45 (8.2%) | 3 (11.1%) | 0 (0.0%) | 110 (5.7%) |
| Stage 2 | 1 (0.2%) | 1 (0.1%) | 9 (1.6%) | 1 (3.7%) | 0 (0.0%) | 12 (0.6%) |
| Stage 3* | 0 (0.0%) | 2 (0.3%) | 8 (1.4%) | 3 (11.1%) | 2 (1.5%) | 15 (0.8%) |
| Any AKI | 23 (4.6%) | 43 (6.0%) | 62 (11.2%) | 7 (25.9%) | 2 (1.5%) | 137 (7.1%) |
| Total | 496 (100.0%) | 717 (100.0%) | 552 (100.0%) | 27 (100.0%) | 130 (100.0%) | 1922 (100.0%) |

† Acute kidney injury at 48 hours defined as an increase from baseline in the cystatin C level by a factor of at least 1.5 to less than 2 (stage 1); an increase from baseline in the cystatin C level by a factor of 2 to 3 (stage 2); or an increase from baseline in the cystatin C level by a factor of more than 3, or the initiation of renal-replacement therapy (stage 3).

*Participants missing Cystatin C levels were assumed to not have AKI except for 2 who had RRT and were classified as stage 3 AKI.

### Supplementary Figure 1: Effect of allocation to rosuvastatin on postoperative cystatin C-defined AKI, overall and according to subgroups

| Subgroup | No. of events/total no. (%) Statin | Placebo | OR (95% CI) |
|---|---|---|---|
| **Age ($\chi^2_1 = 0.94$; p=0.33)** | | | |
| ≤ 60 years | 39/516 (7.6%) | 25/504 (5.0%) | 1.57 (0.93−2.63) |
| >60 years | 49/444 (11.0%) | 24/458 (5.2%) | 2.24 (1.35−3.72) |
| **Sex ($\chi^2_1 = 0.07$; p=0.80)** | | | |
| Male | 66/766 (8.6%) | 35/757 (4.6%) | 1.94 (1.27−2.97) |
| Female | 22/194 (11.3%) | 14/205 (6.8%) | 1.75 (0.87−3.52) |
| **Previous statin use ($\chi^2_1 = 0.4$; p=0.53)** | | | |
| No | 61/639 (9.5%) | 36/630 (5.7%) | 1.74 (1.14−2.67) |
| Yes | 27/321 (8.4%) | 13/332 (3.9%) | 2.25 (1.14−4.45) |
| **Log Troponin I at baseline ($\chi^2_1 = 1.04$; p=0.31)** | | | |
| ≤ 0.04 ng/ml | 71/803 (8.8%) | 42/807 (5.2%) | 1.77 (1.19−2.62) |
| > 0.04 ng/ml | 16/104 (15.4%) | 6/107 (5.6%) | 3.06 (1.15−8.16) |
| **Duration of assigned regimen before surgery ($\chi^2_1 = 0.04$; p=0.85)** | | | |
| ≤ 2 days | 58/554 (10.5%) | 34/577 (5.9%) | 1.87 (1.20−2.90) |
| > 2 days | 30/378 (7.9%) | 15/365 (4.1%) | 2.01 (1.06−3.80) |
| **Surgical technique received ($\chi^2_1 = 0.18$; p=0.68)** | | | |
| On−pump | 61/520 (11.7%) | 33/535 (6.2%) | 2.02 (1.30−3.15) |
| Off−pump | 27/412 (6.6%) | 16/407 (3.9%) | 1.71 (0.91−3.23) |
| **Procedure received ($\chi^2_1 = 0.02$; p=0.88)** | | | |
| CABG | 66/806 (8.2%) | 39/808 (4.8%) | 1.76 (1.17−2.65) |
| AVR | 9/91 (9.9%) | 6/94 (6.4%) | 1.61 (0.55−4.72) |
| **Postoperative glucocorticoid use ($\chi^2_1 = 1.21$; p=0.27)** | | | |
| No | 35/455 (7.7%) | 26/485 (5.4%) | 1.47 (0.87−2.49) |
| Yes | 48/443 (10.8%) | 22/426 (5.2%) | 2.23 (1.32−3.77) |
| **Postoperative NSAID/glucocorticoid use ($\chi^2_1 = 3.07$; p=0.08)** | | | |
| No | 20/321 (6.2%) | 19/341 (5.6%) | 1.13 (0.59−2.15) |
| Yes | 63/577 (10.9%) | 29/570 (5.1%) | 2.29 (1.45−3.61) |
| **AKI risk in those who received CABG and/or AVR ($\chi^2_1 = 2.26$; p=0.13)** | | | |
| Low | 8/307 (2.6%) | 8/310 (2.6%) | 1.01 (0.37−2.73) |
| Medium | 20/311 (6.4%) | 11/307 (3.6%) | 1.85 (0.87−3.93) |
| High | 60/305 (19.7%) | 29/312 (9.3%) | 2.39 (1.49−3.84) |
| **All participants** | **88/960 (9.2%)** | **49/962 (5.1%)** | **1.88 (1.31−2.70)** |

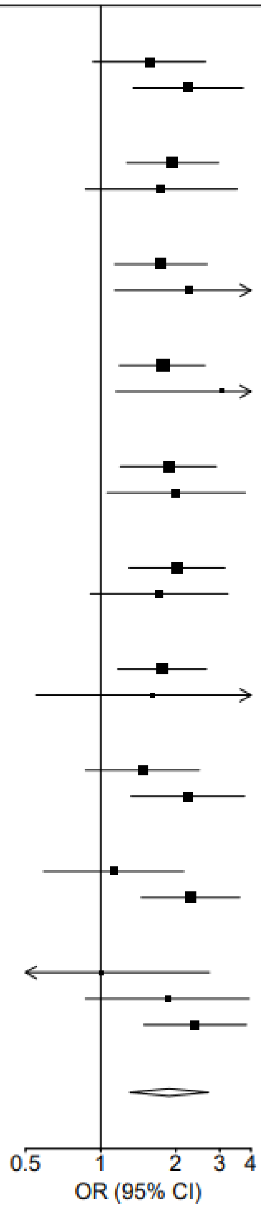


Abbreviations: OR, odds ratio; CI, confidence interval.
128 participants with missing baseline and/or 48hr cystatin C levels were assumed to not have had the event.
The denominator for coronary−artery bypass grafting (CABG) includes only patients who had CABG without aortic−valve replacement, and the denominator for aortic−valve replacement includes only those who had aortic−valve replacement without CABG.
Participants with a missing value for a particular subgroup are not shown (but do contribute to the overall diamond). 101 participants were missing baseline Troponin. 48 participants were scheduled for surgery but did not undergo surgery; these individuals are excluded from the relevant subgroups.
Risk groups were defined using the predicted values from a logistic regression that included age and five variables found to be associated with AKI risk in 1,852 participants that underwent CABG or AVR surgery using an age−adjusted multivariate model that included treatment allocation: baseline NT−pro BNP, baseline KIM−1, procedure (CABG only/AVR only/both), insulin use, and ACE/ARB use.

**Supplementary Figure 2: Effect of rosuvastatin on postoperative creatinine at 48hr according to subgroups**

| Subgroup | Geometric mean (approx. SE) creatinine (mg/dl) Statin | Placebo | % difference in means (95% CI) |
|---|---|---|---|
| **Age ($\chi^2_1$=0.02; p=0.90)** | | | |
| ≤ 60 years | 1.00 (0.01) | 0.97 (0.01) | 3% (0 to 6) |
| >60 years | 1.04 (0.01) | 1.01 (0.01) | 3% (0 to 7) |
| **Sex ($\chi^2_1$=0.18; p=0.67)** | | | |
| Male | 1.02 (0.01) | 0.99 (0.01) | 3% (0 to 5) |
| Female | 1.02 (0.02) | 0.98 (0.02) | 4% (−1 to 10) |
| **Previous statin use ($\chi^2_1$=1.17; p=0.28)** | | | |
| No | 1.02 (0.01) | 0.99 (0.01) | 4% (1 to 7) |
| Yes | 1.01 (0.01) | 1.00 (0.01) | 1% (−2 to 5) |
| **Log Troponin I at baseline ($\chi^2_1$=2.95; p=0.09)** | | | |
| ≤ 0.04 ng/ml | 1.02 (0.01) | 0.99 (0.01) | 3% (0 to 5) |
| > 0.04 ng/ml | 1.03 (0.02) | 0.94 (0.02) | 9% (2 to 16) |
| **Duration of assigned regimen before surgery ($\chi^2_1$=0.30; p=0.58)** | | | |
| ≤ 2 days | 1.01 (0.01) | 0.99 (0.01) | 3% (0 to 6) |
| > 2 days | 1.04 (0.01) | 1.00 (0.01) | 4% (0 to 8) |
| **Surgical technique received ($\chi^2_1$=0.51; p=0.47)** | | | |
| On−pump | 1.03 (0.01) | 0.99 (0.01) | 4% (1 to 7) |
| Off−pump | 1.01 (0.01) | 0.99 (0.01) | 2% (−1 to 6) |
| **Procedure received ($\chi^2_1$=0.03; p=0.86)** | | | |
| CABG only | 1.01 (0.01) | 0.99 (0.01) | 3% (0 to 5) |
| AVR only | 1.00 (0.03) | 0.98 (0.02) | 2% (−5 to 9) |
| **Postoperative glucocorticoid use ($\chi^2_1$=0.02; p=0.89)** | | | |
| No | 1.02 (0.01) | 0.99 (0.01) | 3% (0 to 7) |
| Yes | 1.02 (0.01) | 0.99 (0.01) | 3% (0 to 6) |
| **Postoperative NSAID/glucocorticoid use ($\chi^2_1$=0.11; p=0.74)** | | | |
| No | 1.02 (0.01) | 0.98 (0.01) | 4% (0 to 8) |
| Yes | 1.02 (0.01) | 1.00 (0.01) | 3% (0 to 6) |
| **All participants** | **1.02 (0.01)** | **0.99 (0.01)** | **3% (1 to 5)** |

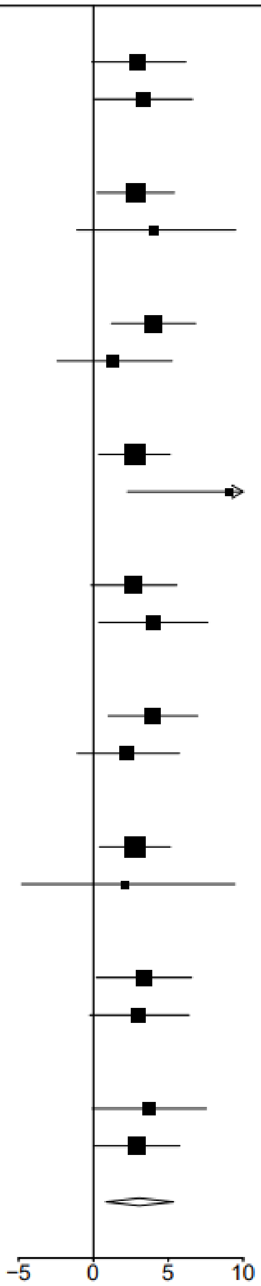


Abbreviations: Ln, natural logarithm; CI, confidence interval; SE, standard error.
Estimates were derived from analysis of covariance with adjustment for the baseline value. Geometric means with approximate SE are presented.
P−values were from a heterogeneity test comparing the difference in means.
Missing biomarker data for 144 participants were imputed with the use of multiple imputation.
Participants with a missing value for a particular subgroup are not shown (but do contribute to the overall diamond).

**Supplementary Figure 3: Effect of rosuvastatin on postoperative cystatin C at 48hr according to subgroups**

| Subgroup | Geometric mean (approx. SE) cystatin C (mg/L) Statin | Placebo | % difference in means (95% CI) |
|---|---|---|---|
| **Age ($\chi^2_1$=1.00; p=0.32)** | | | |
| ≤ 60 years | 1.05 (0.01) | 1.00 (0.01) | 4% (1 to 7) |
| >60 years | 1.10 (0.01) | 1.03 (0.01) | 7% (3 to 10) |
| **Sex ($\chi^2_1$=0.06; p=0.80)** | | | |
| Male | 1.07 (0.01) | 1.02 (0.01) | 5% (3 to 8) |
| Female | 1.08 (0.02) | 1.02 (0.02) | 6% (1 to 11) |
| **Previous statin use ($\chi^2_1$=0.18; p=0.67)** | | | |
| No | 1.07 (0.01) | 1.01 (0.01) | 6% (3 to 8) |
| Yes | 1.07 (0.01) | 1.02 (0.01) | 5% (1 to 9) |
| **Log Troponin I at baseline ($\chi^2_1$=1.90; p=0.17)** | | | |
| ≤ 0.04 ng/ml | 1.07 (0.01) | 1.02 (0.01) | 5% (3 to 7) |
| > 0.04 ng/ml | 1.08 (0.02) | 0.98 (0.02) | 10% (3 to 16) |
| **Duration of assigned regimen before surgery ($\chi^2_1$=0.38; p=0.54)** | | | |
| ≤ 2 days | 1.08 (0.01) | 1.02 (0.01) | 6% (3 to 9) |
| > 2 days | 1.06 (0.01) | 1.02 (0.01) | 5% (1 to 8) |
| **Surgical technique ($\chi^2_1$=0.25; p=0.62)** | | | |
| On−pump | 1.09 (0.01) | 1.03 (0.01) | 6% (3 to 9) |
| Off−pump | 1.04 (0.01) | 0.99 (0.01) | 5% (2 to 8) |
| **Procedure ($\chi^2_1$=1.15; p=0.28)** | | | |
| CABG only | 1.06 (0.01) | 1.01 (0.01) | 4% (2 to 7) |
| AVR only | 1.10 (0.03) | 1.02 (0.02) | 8% (2 to 15) |
| **Postoperative glucocorticoid use ($\chi^2_1$=0.96; p=0.33)** | | | |
| No | 1.07 (0.01) | 1.00 (0.01) | 6% (3 to 9) |
| Yes | 1.08 (0.01) | 1.04 (0.01) | 4% (1 to 7) |
| **Postoperative NSAID/glucocorticoid use ($\chi^2_1$=0.14; p=0.71)** | | | |
| No | 1.06 (0.01) | 1.00 (0.01) | 6% (2 to 10) |
| Yes | 1.08 (0.01) | 1.03 (0.01) | 5% (2 to 8) |
| **All participants** | **1.07 (0.01)** | **1.02 (0.01)** | **5% (3 to 8)** |

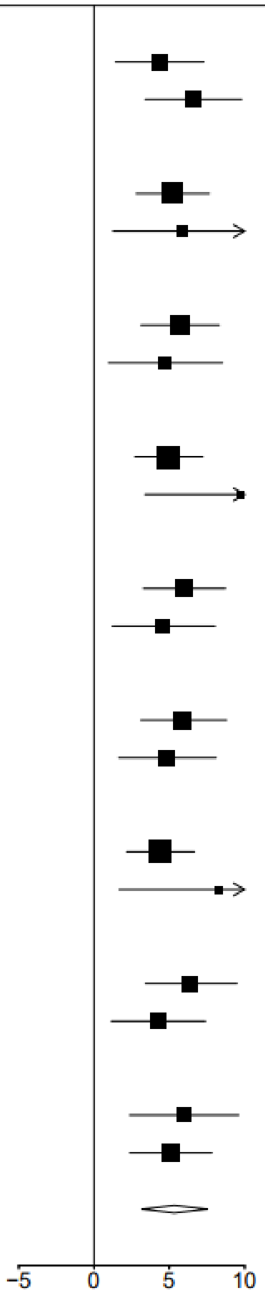


Abbreviations: Ln, natural logarithm; CI, confidence interval; SE, standard error.
Estimates were derived from analysis of covariance with adjustment for the baseline value. Geometric means with approximate SE are presented.
P−values were from a heterogeneity test comparing the difference in means.
Missing biomarker data for 130 participants were imputed with the use of multiple imputation.
Participants with a missing value for a particular subgroup are not shown (but do contribute to the overall diamond).
